# Prospective neuroimaging and neuropsychological evaluation in adults with newly diagnosed focal epilepsy

**DOI:** 10.1101/2024.05.14.24307267

**Authors:** Christophe de Bezenac, Nicola Leek, Guleed Adan, Ahmad Ali, Rajiv Mohanraj, Shubhabrata Biswas, Ronan Mcginty, Kieran Murphy, Helen Malone, Gus Baker, Perry Moore, Anthony G Marson, Simon S. Keller

## Abstract

**Objective:** There are few prospective longitudinal studies in patients with newly diagnosed epilepsy (NDE) despite that this is a key time point to understand the underlying biology of epilepsy and to identify potential interventions and biomarkers for seizure and cognitive outcomes. Here we have performed a prospective combined neuroimaging and neuropsychological study in a cohort of patients with focal NDE and healthy controls.

**Methods:** We recruited 104 patients with NDE and 45 healthy controls for research-grade 3 Tesla MRI (diagnostic and structural imaging, diffusional kurtosis imaging, resting-state functional MRI, task-based functional MRI), EEG, comprehensive neuropsychological, and blood biomarker investigations. We report here on the baseline clinical, neuroradiological, MRI morphometric, and neuropsychological findings in this cohort.

**Results:** 38% of patients had unremarkable MRI features, 12% had lesions of known significance in epilepsy, 49% with abnormalities of unknown significance in epilepsy, and 23% with incidental findings. In comparison, 56% of controls had unremarkable MRI features, 7% had lesions of known significance in epilepsy, 33% with abnormalities of unknown significance in epilepsy, and 16% had incidental findings. Patients had a higher incidence of white matter hyperintensities compared to controls. Reduced bihemispheric frontal lobe cortical thickness and thalamic volumes were observed in patients with moderate effect sizes. Patients scored significantly lower on tasks of executive function, processing speed, and visual, delayed, and immediate memory, and significantly higher on depression and anxiety assessments compared to controls. Patient neuropsychological performance was related to various brain morphometric features.

**Significance:** People with adult focal NDE have a greater proportion of MRI-positive findings than previously reported. Subtle white matter lesions may represent an important diagnostic criterion and have a pathophysiological basis in focal epilepsy. Morphometric and neuropsychological alterations are present at the point of diagnosis of epilepsy, which suggests that brain and cognitive changes are not exclusively due to the deleterious impact of chronic epilepsy.

## Introduction

The underlying biology of human epilepsy is significantly understudied from the point of diagnosis. The translation of what we understand in longstanding epilepsy to people with a new diagnosis of epilepsy is confounded by several factors, including the chronic effects of seizures and anti-seizure medication.^1^ At present we have very limited ability to stratify newly diagnosed patients for important outcomes such as seizure control and cognitive dysfunction, which diminishes our ability to individualise patient management and counselling.^2, 3^ A detailed prospective study using novel imaging techniques in newly diagnosed epilepsy (NDE) is required to identify important biomarkers associated with biological processes underpinning the disease, seizure outcome and comorbidities. However, patients have only rarely been prospectively studied from the time of diagnosis, particularly using combined research grade neuroimaging and cognitive investigations, despite this being a key point in time to understand the underlying biology of epilepsy and to identify potential interventions and biomarkers for seizure and cognitive outcomes.^1, 4^

Many people with epilepsy are cognitively impaired at the time of diagnosis. Drug naïve patients with NDE show significant impairments in memory, sustained attention, executive functioning, mental flexibility and psychomotor speed relative to healthy volunteers.^5–10^ One 12-month follow-up study revealed that performance on some of these cognitive domains further deteriorate.^11^ Cognitive deficits – which along with spontaneous seizures contribute to impaired quality of life in epilepsy^12^ - are therefore not necessarily a result of the chronicity of the disorder, despite that cognitive impairment is likely to be exacerbated by ongoing seizures and the burden of epilepsy.^13^ However, there are currently no insights from existing neuroimaging investigations as to the underlying mechanisms of cognitive dysfunction at the point of epilepsy diagnosis. Given that impairments in brain structural and functional networks – facets of brain organisation only amenable to investigation using research grade neuroimaging methods – are becoming increasingly recognised as important markers of seizure phenotypes, treatment outcomes and cognitive impairment in refractory epilepsy,^14–16^ it is crucial that brain networks and other aspects of brain architecture and function not amenable to investigation using standard clinical MRI methods be prospectively studied from the earliest point in human epilepsy using advanced imaging methods.

In order to address this research gap, we have designed a research programme that recruited patients with focal NDE for advanced MRI, resting-state EEG, detailed neuropsychological evaluation, and analysis of blood samples with six-monthly follow up to determine patterns of seizure relapse and remission.^17^ The primary objectives of this programme of research is to use the detailed and high fidelity neuroimaging, neurophysiological, neuropsychological and biological data to understand the mechanisms of, and stratify patients according to cognitive phenotypes and anti-seizure medication treatment outcomes. The goal of this paper is to describe the cohort and present baseline data. We sought to classify neuroradiological findings based on and extending previously published criteria in NDE,^18^ present basic quantitative MRI findings, and summarise neuropsychological results, exploring interactions between the clinical, diagnostic, imaging, and cognitive data.

## Methods

This study was approved by the Northwest Liverpool East Research Ethics Committee (19/NW/0384) through the Integrated Research Application System (Project ID 260623). The research is sponsored by the University of Liverpool (UoL001449) and UK Health Research Authority approval was provided on 22^nd^ August 2019. Recruitment of the first participant was 24^th^ August 2019.

### Participants

Patients with focal NDE were recruited from the Walton Centre NHS Foundation Trust and Salford Royal (now Northern Care Alliance) NHS Foundation Trust in the UK. Standardised EEG investigations were performed at the respective NHS Trusts. All patients received MRI, neuropsychological evaluation, and blood extraction and storage at the University of Liverpool. The inclusion criteria for patients included diagnosis of focal onset epilepsy by an experienced neurologist based on seizure semiology at the two NHS Trusts, between and including the ages 16-70 years, and all investigations to be performed within three months of diagnosis. We chose not to recruit drug-naïve patients as this would have substantially limited the number of eligible patients within the timeframe of the study. We did not anticipate any significant deleterious impact of brief use of anti-seizure medication on imaging or cognitive data. Patient exclusion criteria included non-epileptic seizures, primary generalised seizures, single seizures, provoked seizures only (e.g., alcohol), known inflammatory neurological condition (specifically multiple sclerosis or sarcoidosis), acute symptomatic seizures (e.g., acute brain haemorrhage or brain injury), progressive neurological disease (e.g., known brain tumour), previous neurosurgery, concomitant infection, and any other significant comorbidity (at physicians’ discretion). As done previously,^19^ seizures were classified as focal without impairment of awareness (including seizures with observable motor or autonomic phenomena and seizures with subjective sensory and psychic phenomena), focal with impairment of awareness, focal with evolution to bilateral tonic-clonic, or unclassifiable.

We additionally recruited healthy controls aged, sex, and education matched to patients with no history of neurological or psychiatric illness or disease. All controls underwent the same investigations as patients. Based on power calculations we intended to recruit 107 patients and 48 controls for MRI, EEG, neuropsychological assessment, and blood sampling.^17^ The calculation was based on the number of patients expected to be seizure-free and seizure-active two years after recruitment, the number of MRI-negative/lesional cases, and 10% attrition to follow up. All patients underwent follow-up after neuroimaging and neuropsychological assessment at six-month intervals for two years to capture the number of seizures experienced in the preceding six months and current medications and dosage. Blood was collected for analysis for all participants in lithium-heparin bottles or serum separator tubes (maximum of 72 mL of blood (3×9 mL vials)) by a healthcare professional trained in phlebotomy. Blood samples have been stored in the University of Liverpool Biobank and will be subject to investigation of peripherally circulating cytokines and markers of cell damage in due course.

### Neuroimaging

All participants underwent MRI scanning at the Liverpool Magnetic Resonance Imaging Centre (LiMRIC) at the University of Liverpool using a 3 T Siemens Magnetom Prisma scanner using a 32-channel head coil. The MRI protocol consisted of structural sequences for screening of incidental findings and identification of lesions, a 3D T1-weighted scan, diffusional kurtosis imaging, resting-state functional MRI, and a verbal memory functional MRI paradigm adapted from a previous study.^20^ Further information on each MRI sequence is provided in Supplementary Table 1.

A consultant neuroradiologist (SB) with expertise in epilepsy neuroimaging reviewed all patient and healthy control scans for lesions and incidental findings. Participant age and new diagnosis of epilepsy was known to the neuroradiologist, but no other information was provided. Neuroradiological features were classified as normal, incidental, or abnormal, and abnormal further divided into likely epilepsy-related or unknown relationship to epilepsy, as previously done.^19^ Epilepsy-related lesions were ascribed to abnormalities with a previously reported potential for epileptogenicity including malformations of cortical development, hippocampal sclerosis, foreign tissue lesions, and focal encephalomalacia or gliosis. Abnormalities characterised as unknown relationship to epilepsy included but were not restricted to diffuse white matter changes, diffuse brain atrophy, asymmetric hippocampi and amygdlae with no clear evidence of unilateral sclerosis, and historical cerebellar infarcts. Unlike previous work,^19^ we classified supratentorial arachnoid cysts as unknown relationship to epilepsy rather than incidental findings unless they had been ruled out to be epileptogenic by EEG and seizures confidently localised elsewhere. Possible causal relationship between epilepsy and arachnoid cysts has been suggested in various publications.^21–23^ All remaining cysts were classified as incidental.

For the purposes of this investigation, we performed volumetry of subcortical, cerebral lobar, and cerebellar regions to determine whether patients showed evidence of gross brain atrophy compared to controls, and whether volumetric alterations were associated with neuropsychological changes. We used FreeSurfer (version 7.1.1) to extract cortical thickness and subcortical volume for brain regions. FreeSurfer is an open source neuroimage analysis suite available at https://surfer.nmr.mgh.harvard.edu/. The recon-all pipeline with default settings was used, which included stages such as motion correction, averaging of multiple T1 images, removal of non-brain tissue using a watershed/surface deformation procedure, and segmentation of white matter and subcortical grey matter in Talairach space. FreeSurfer segmentation labels were derived from probabilistic information automatically estimated from expert segmentations of 40 adult brain images.^24^

Further technical details of the FreeSurfer process can be found in previous publications.^24, 25^ A trained technician visually examined all processed images. No participants were excluded from the study due to artifacts, movement errors, or structural anomalies. The measurements of interest comprised cortical mean cortical thickness, and cortical volume parcellated into 34 bilateral regions of interest (ROIs) as per Desikan et al.,^26^ along with total brain volume.

### EEG

All patients and controls underwent conventional diagnostic EEG recordings using the international 10-20 system. Each EEG acquisition was evaluated by an expert neurophysiologist for evidence of inter-ictal epileptiform activity. Each EEG acquisition included resting-state data, which was identified by a trained clinical EEG technician. This data will be subject to physiological network analysis in due course.^17^

### Neuropsychology

All participants underwent computerised neuropsychological assessment using a tailored battery that targeted cognitive domains previously shown to be impacted in patients with NDE.^5, 11^ The battery included components from the Wechsler Memory Scale Fourth Edition (WMS-IV),^27^ Wechsler Adult Intelligence Scale Fourth Edition (WAIS-IV),^11, 27^ Delis-Kaplan Executive Function System (D-KEFS),^28, 29^ Patient Health Questionnaire 9 (PHQ-9),^30^ Generalised Anxiety Disorder 7 (GAD-7),^31^ Perceived Deficits Questionnaire (PDQ),^32^ and Quality Of Life In Epilepsy (QOLIE) scale.^33^ These assessment tools were used to evaluate auditory memory through story recall and recall of verbal pairs (WMS-IV), visual memory through the visual reproduction of drawings and the recall of content and spatial features (WMS-IV), auditory working memory (WAIS-IV), attention span and executive control (WAIS-IV), digit span and arithmetic (WAIS-IV), processing and psychomotor speed (WAIS-IV), coding and symbol search (WAIS-IV), finger tapping and visual reaction time (WAIS-IV), verbal fluency and executive functioning (the Stroop task) (D-KEFS), mood including depression (PHQ-9) and anxiety (GAD-7), perceived cognitive impairment (PDQ), and quality of life (QOLIE). Summary indices were computed for memory (Auditory.Memory, Delayed.Memory, Immediate.Memory, Visual.Memory) and cognition (Working.Memory, Processing.Speed, Executive.Function). Other represented domains included mood (Depression.PHQ9, Anxiety.GAD7) and motor function (Finger.Tapping.RH, Finger.Tapping.LH, Visual.RT.M, Visual.RT.SD),

### Statistical analysis

Data analysis was performed using R (version 4.2.0, https://www.R-project.org/). Tables with associated summary statistics were generated with the R package arsenal (version 3.6.3). Analysis of variance test (ANOVA) was used for continuous variables, chi-square goodness of fit test was used for categorical or factor variables, and trend test was used for equal distribution of ordered factor variables.

For each neuropsychological feature, we employed a participant-specific z-score computation through a resampling approach. This method involved determining the mean and standard deviation of feature scores for 100 random samples of 10 healthy control participants (ensuring that participants for whom the z-score was being computed was excluded from these samples in the case of controls). The difference between a participant’s feature score and the mean of the 100 computed mean values was divided by the mean of the 100 standard deviation values [z = (score - mean(100 x mean(scores for 10 controls))) / mean(100 x sd(scores for 10 controls))]. This process resulted in a z-score per feature for both patients and controls.

A z-score threshold greater than two (z-score > 2) was applied for features where we anticipated patients to exhibit higher scores (Depression.PHQ9, Anxiety.GAD7, Visual.RT.M, Visual.RT.SD). Conversely, a threshold smaller than two (z-score < 2) was used for features where we expected patients to perform worse based on previous work (Finger.Tapping.RH, Finger.Tapping.LH, Auditory.Memory, Delayed.Memory, Immediate.Memory, Visual.Memory, Working.Memory, Processing.Speed, Executive.Function).^5–11^

We assessed structural brain difference between patients with NDE and healthy control subjects using multiple linear regressions (lm function in R). Brain volume was used for subcortical structures and mean thickness for cortical regions. A binary diagnosis indicator (0 = healthy control, 1 = person with epilepsy) served as the predictor of interest, and the volume or thickness of a designated brain region constituted the outcome measure. Effect size estimates across all brain regions were computed as Cohen’s d, adjusting for age, sex, and intracranial volume (ICV) (to control for the association between volume with head size). To address multiple comparisons, we applied a false discovery rate (FDR) correction at q=0.05.

## Results

### Cohort

The Covid pandemic had a significant impact on recruitment. Given the closure of imaging facilities, prioritisation of covid-related healthcare, and repurposing of clinical staff, recruitment was delayed by 16 months (Supplementary Figure 1). We recruited 104 patients with focal NDE and 45 healthy controls, three patients and three controls short of our target. There were no significant differences between patients and controls in age, sex, or education, although there was a trend for controls to be older than patients (Table 1). Most patients did not have a familial history of epilepsy, fewer than 10 seizures at the time of diagnosis, did not have nocturnal seizures, experienced convulsive seizures with loss of awareness, had seizures preceded by an aura, and no history of febrile seizures, brain infection, head injury, or birth complications (Table 1). 22.1% of patients reported to have experienced ten or more seizures prior to diagnosis, and 4.2% reported over 100 seizure events.

**Table 1.** Summary of demographic and clinical data.

|  | Control (N=45) | Patient (N=104) | Total (N=149) | p value |
| --- | --- | --- | --- | --- |
| **Age** |  |  |  | 0.052 |
| Mean (SD) | 40.752 (15.745) | 35.777 (13.526) | 37.280 (14.362) |  |
| Range | 17.205 - 68.371 | 16.657 - 69.228 | 16.657 - 69.228 |  |
| **Sex** |  |  |  | 0.868 |
| Male | 24 (53.3%) | 57 (54.8%) | 81 (54.4%) |  |
| Female | 21 (46.7%) | 47 (45.2%) | 68 (45.6%) |  |
| **Completed high school** |  |  |  | 0.765 |
| Yes | 28 (62.2%) | 62 (59.6%) | 90 (60.4%) |  |
| No | 17 (37.8%) | 42 (40.4%) | 59 (39.6%) |  |
| **Ethnicity** |  |  |  | 0.697 |
| White British | 40 (88.9%) | 94 (90.4%) | 134 (89.9%) |  |
| White other | 1 (2.2%) | 4 (3.8%) | 5 (3.4%) |  |
| Multiple | 1 (2.2%) | 3 (2.9%) | 4 (2.7%) |  |
| African | 1 (2.2%) | 2 (1.9%) | 3 (2.0%) |  |
| Asian | 2 (4.4%) | 1 (1.0%) | 3 (2.0%) |  |
| **Family history of epilepsy** |  |  |  | < 0.001 |
| No | 45 (100.0%) | 80 (76.9%) | 125 (83.9%) |  |
| Yes | 0 (0.0%) | 24 (23.1%) | 24 (16.1%) |  |
| **Number of seizures** |  |  |  |  |
| N-Miss |  | 9 |  |  |
| Five or less |  | 61 (64.2%) |  |  |
| More than five |  | 34 (35.8%) |  |  |
| **Nocturnal seizures** |  |  |  |  |
| N-Miss |  | 0 |  |  |
| Yes |  | 80 (76.9%) |  |  |
| No |  | 19 (18.3%) |  |  |
| Sometimes |  | 5 (4.8%) |  |  |
| **Convulsive seizures** |  |  |  |  |
| N-Miss |  | 0 |  |  |
| Yes |  | 73 (70.2%) |  |  |
| No |  | 24 (23.1%) |  |  |
| Unsure |  | 7 (6.7%) |  |  |
| **Loss of awareness** |  |  |  |  |
| N-Miss |  | 0 |  |  |
| Yes |  | 61 (58.7%) |  |  |
| No |  | 19 (18.3%) |  |  |
| Sometimes |  | 5 (4.8%) |  |  |
| Unsure |  | 19 (18.3%) |  |  |
| **Seizures with aura** |  |  |  |  |
| N-Miss |  | 0 |  |  |
| Yes |  | 56 (53.8%) |  |  |
| No |  | 28 (26.9%) |  |  |
| Sometimes |  | 1 (1.0%) |  |  |
| Unsure |  | 19 (18.3%) |  |  |
| **Febrile seizures** |  |  |  |  |
| N-Miss |  | 0 |  |  |
| Yes |  | 11 (10.6%) |  |  |
| No |  | 88 (84.6%) |  |  |
| Unsure |  | 5 (4.8%) |  |  |
| **History of brain infection** |  |  |  |  |
| N-Miss |  | 0 |  |  |
| Yes |  | 1 (1.0%) |  |  |
| No |  | 100 (96.2%) |  |  |
| Unsure |  | 3 (2.9%) |  |  |
| **History of head injury** |  |  |  |  |
| N-Miss |  | 0 |  |  |
| Yes |  | 19 (18.3%) |  |  |
| No |  | 84 (80.8%) |  |  |
| Unsure |  | 1 (1.0%) |  |  |
| **Birth complications** |  |  |  |  |
| N-Miss |  | 0 |  |  |
| Yes |  | 21 (20.2%) |  |  |
| No |  | 79 (76.0%) |  |  |
| Unsure |  | 4 (3.8%) |  |  |
| **Outcome at 6 months** |  |  |  |  |
| N-Miss |  | 10 |  |  |
| PS |  | 41 (43.6%) |  |  |
| SF |  | 53 (56.4%) |  |  |
| **Outcome at 12 months** |  |  |  |  |
| N-Miss |  | 27 |  |  |
| PS |  | 32 (41.6%) |  |  |
| SF |  | 45 (58.4%) |  |  |
| **Outcome at 18 months** |  |  |  |  |
| N-Miss |  | 36 |  |  |
| PS |  | 22 (32.4%) |  |  |
| SF |  | 46 (67.6%) |  |  |
| **Outcome at 24 months** |  |  |  |  |
| N-Miss |  | 52 |  |  |
| PS |  | 22 (42.3%) |  |  |
| SF |  | 30 (57.7%) |  |  |

Figure 1 and Table 1 shows the recorded post-treatment outcome for patients in whom this data was available at the time of writing. There were 56.7%, 57.5%, 66.7% and 61.7% patients who did not experience a seizure within the preceding six months at month 6, 12, 18, and 24, respectively. Many patients did not have consistent outcomes across each 6-month period (Figure 1). Of the 83 patients with 24 months outcome to date (and ignoring missing data), only 37% and 18% were consistently seizure free or experienced seizures across every 6-month period, respectively.

**Figure 1.**
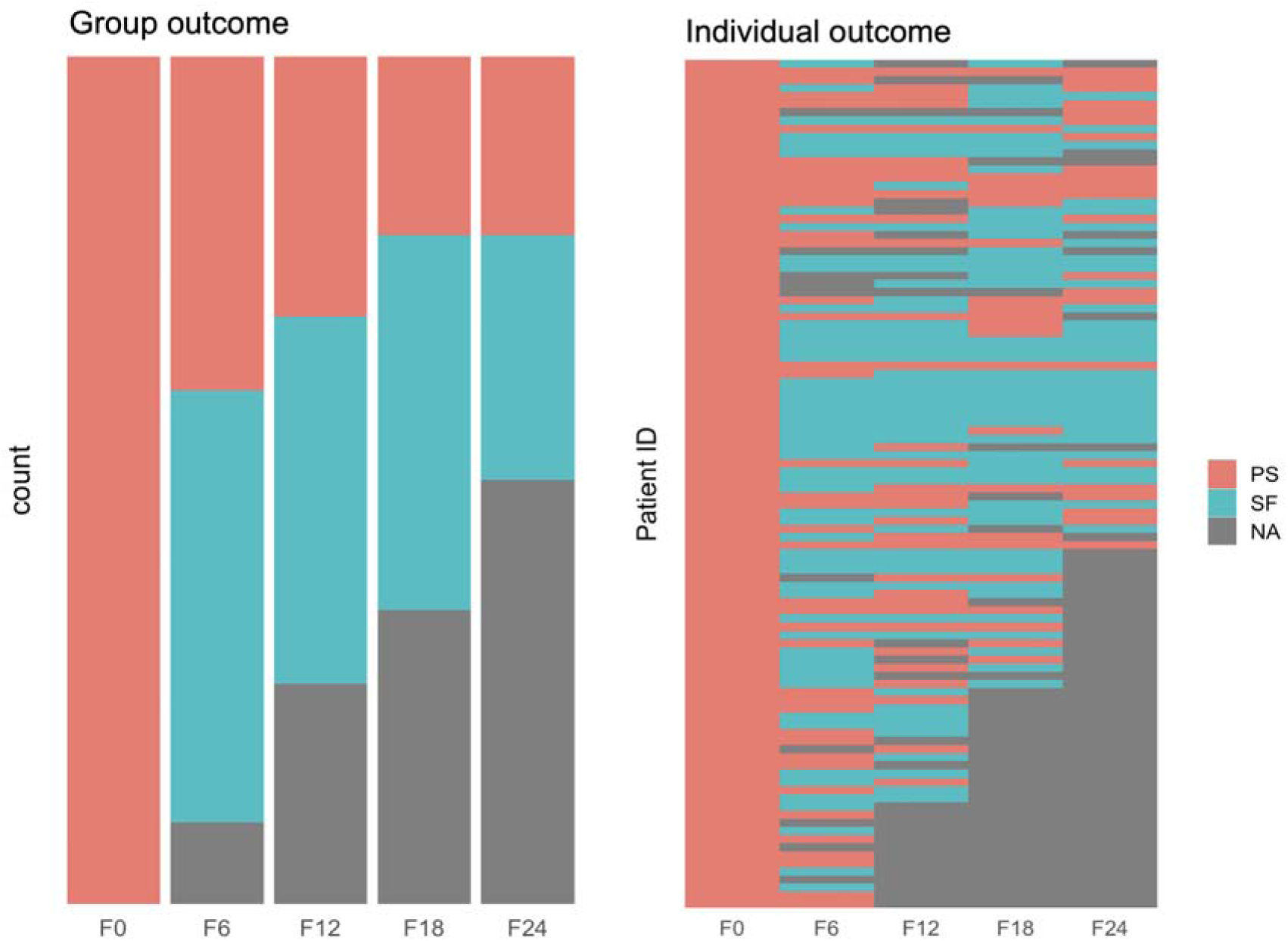
Patient outcome over time. The proportion of patients with NDE that have persistent seizures (PS) and those that are seizure free (SF) for 0 (time of testing), 6-, 12-, 18-, and 24-months follow-ups (left). Individual-level outcomes over time (right). Grey indicates data missing or not-yet-acquired.

### Neuroradiological findings

Table 2 presents a summary of the neuroradiological findings. Sixty one percent of patients had MRI reports classified as abnormal, compared to 44% of controls (p = 0.053). There were 38% of patients with unremarkable MRI features, 12% with lesions of known significance in epilepsy, 49% with abnormalities of unknown significance in epilepsy, and 23% with incidental findings. In comparison, 56% of controls had unremarkable MRI features, 7% had lesions of known significance in epilepsy, 33% with abnormalities of unknown significance in epilepsy, and 16% had incidental findings. Figure 2 provides examples of MRI detected lesions classified as epilepsy related, unknown relevance to epilepsy, and incidental.

**Figure 2.**
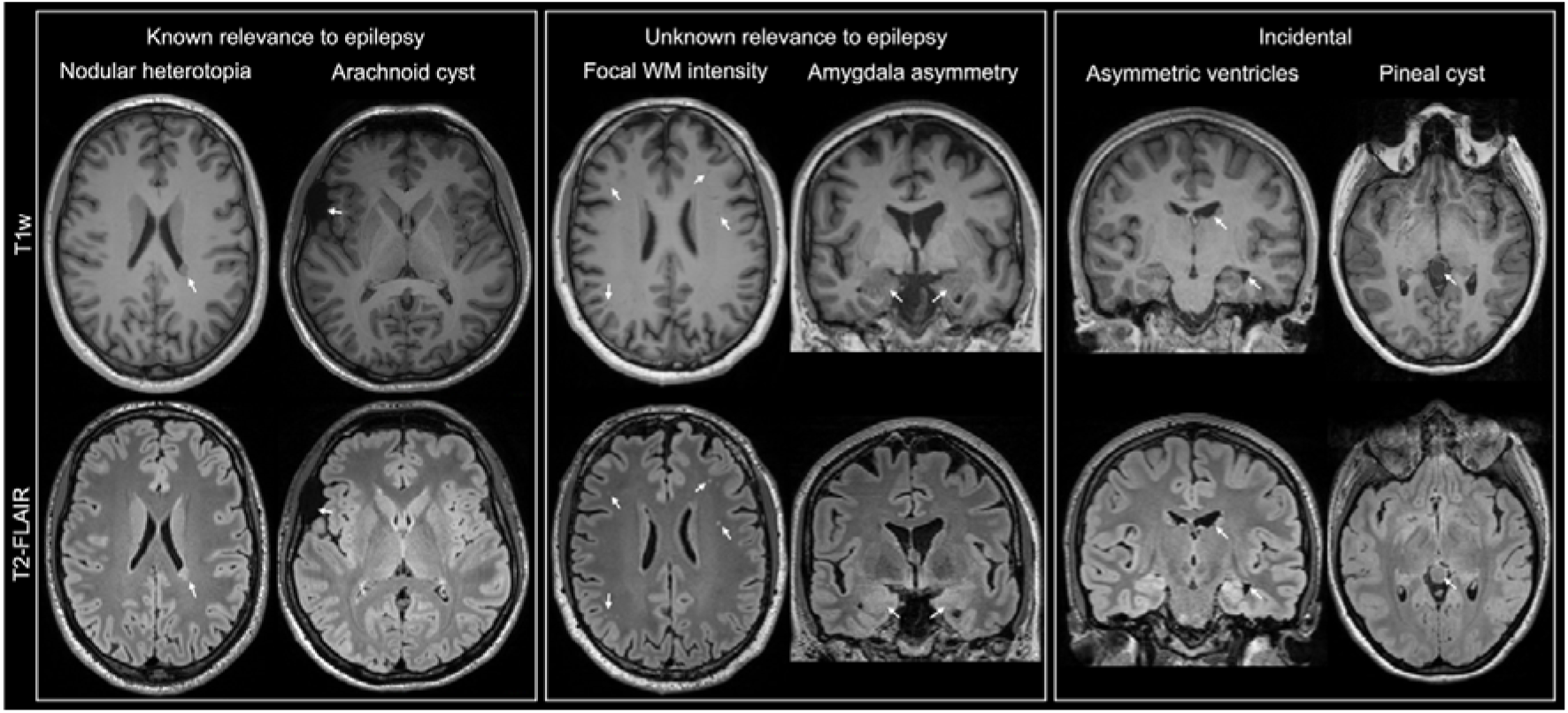
Exemplar MRI-positive cases across the three lesional categories. Known relevance to epilepsy: Nodular heterotopia left supra-trigonal periventricular region (left) and arachnoid cyst overlying anterior aspect of the right sylvian fissure, abutting the inferior frontal gyrus superiorly and the temporal pole inferiorly (right). Unknown relevance to epilepsy: multiple focal white matter hypointensities (T1w) and hyperintensities (T2-FLAIR) (left) and amygdala asymmetry (right; note additional ventricular asymmetry). Incidental: asymmetric lateral ventricles, including frontal and temporal horns (left) and pineal cyst (right).

**Table 2.** Neuroradiological and neurophysiological findings.

|  | Control (N=45) | Patient (N=104) | Total (N=149) | p value |
| --- | --- | --- | --- | --- |
| **MRI.Report.Conclusion** |  |  |  | 0.053 |
| Abnormal | 20 (44.4%) | 64 (61.5%) | 84 (56.4%) |  |
| Normal | 25 (55.6%) | 40 (38.5%) | 65 (43.6%) |  |
| **EEG.Conclusion** |  |  |  | 0.638 |
| N-Miss | 4 | 8 | 12 |  |
| Abnormal | 8 (19.5%) | 13 (13.5%) | 21 (15.3%) |  |
| Unknown | 3 (7.3%) | 6 (6.2%) | 9 (6.6%) |  |
| Normal | 30 (73.2%) | 77 (80.2%) | 107 (78.1%) |  |
| **Epilepsy.Related.Single.Lesion** |  |  |  | 0.453 |
| Yes | 3 (6.7%) | 11 (10.6%) | 14 (9.4%) |  |
| No | 42 (93.3%) | 93 (89.4%) | 135 (90.6%) |  |
| **Epilepsy.Related.Cavernoma** |  |  |  | 0.127 |
| Yes | 1 (2.2%) | 0 (0.0%) | 1 (0.7%) |  |
| No | 44 (97.8%) | 104 (100.0%) | 148 (99.3%) |  |
| **Epilepsy.Related.Neoplasm** |  |  |  | 0.182 |
| Yes | 0 (0.0%) | 4 (3.8%) | 4 (2.7%) |  |
| No | 45 (100.0%) | 100 (96.2%) | 145 (97.3%) |  |
| **Epilepsy.Related.Malformations.Of.Cortical.Development** |  |  |  | 0.250 |
| Yes | 0 (0.0%) | 3 (2.9%) | 3 (2.0%) |  |
| No | 45 (100.0%) | 101 (97.1%) | 146 (98.0%) |  |
| **Epilepsy.Related.Heterotopia** |  |  |  | 0.349 |
| Yes | 0 (0.0%) | 2 (1.9%) | 2 (1.3%) |  |
| No | 45 (100.0%) | 102 (98.1%) | 147 (98.7%) |  |
| **Epilepsy.Related.Supratentorial.Old.Stroke** |  |  |  | 0.509 |
| Yes | 0 (0.0%) | 1 (1.0%) | 1 (0.7%) |  |
| No | 45 (100.0%) | 103 (99.0%) | 148 (99.3%) |  |
| **Epilepsy.Related.Supratentorial.Encephalomalacia** |  |  |  | 0.627 |
| Yes | 2 (4.4%) | 3 (2.9%) | 5 (3.4%) |  |
| No | 43 (95.6%) | 101 (97.1%) | 144 (96.6%) |  |
| **Unknown.Single.Lesion** |  |  |  | 0.076 |
| Yes | 15 (33.3%) | 51 (49.0%) | 66 (44.3%) |  |
| No | 30 (66.7%) | 53 (51.0%) | 83 (55.7%) |  |
| **Unknown.Multiple.Lesions** |  |  |  | 0.027 |
| Yes | 1 (2.2%) | 15 (14.4%) | 16 (10.7%) |  |
| No | 44 (97.8%) | 89 (85.6%) | 133 (89.3%) |  |
| **Unknown.Other** |  |  |  | 0.349 |
| Yes | 0 (0.0%) | 2 (1.9%) | 2 (1.3%) |  |
| No | 45 (100.0%) | 102 (98.1%) | 147 (98.7%) |  |
| **Unknown.Other.Supratentorial.Hyperintensity** |  |  |  | 0.818 |
| Yes | 1 (2.2%) | 3 (2.9%) | 4 (2.7%) |  |
| No | 44 (97.8%) | 101 (97.1%) | 145 (97.3%) |  |
| **Unknown.Arachnoid.Cyst** |  |  |  | 0.865 |
| Yes | 2 (4.4%) | 4 (3.8%) | 6 (4.0%) |  |
| No | 43 (95.6%) | 100 (96.2%) | 143 (96.0%) |  |
| **Unknown.Other.Supratentorial.Neoplasm** |  |  |  | 0.509 |
| Yes | 0 (0.0%) | 1 (1.0%) | 1 (0.7%) |  |
| No | 45 (100.0%) | 103 (99.0%) | 148 (99.3%) |  |
| **Unknown.Asymmetric.Amygdala** |  |  |  | 0.135 |
| Yes | 0 (0.0%) | 5 (4.8%) | 5 (3.4%) |  |
| No | 45 (100.0%) | 99 (95.2%) | 144 (96.6%) |  |
| **Unknown.Asymmetric.Hippocampus** |  |  |  | 0.349 |
| Yes | 0 (0.0%) | 2 (1.9%) | 2 (1.3%) |  |
| No | 45 (100.0%) | 102 (98.1%) | 147 (98.7%) |  |
| **Unknown.Hyperintensity.In.Amygdala.Or.Hippocampus** |  |  |  | 0.182 |
| Yes | 0 (0.0%) | 4 (3.8%) | 4 (2.7%) |  |
| No | 45 (100.0%) | 100 (96.2%) | 145 (97.3%) |  |
| **Unknown.Cortical.Atrophy** |  |  |  | 0.865 |
| Yes | 2 (4.4%) | 4 (3.8%) | 6 (4.0%) |  |
| No | 43 (95.6%) | 100 (96.2%) | 143 (96.0%) |  |
| **Unknown.White.Matter.Hyperintensities** |  |  |  | 0.078 |
| Yes | 11 (24.4%) | 41 (39.4%) | 52 (34.9%) |  |
| No | 34 (75.6%) | 63 (60.6%) | 97 (65.1%) |  |
| **Incidental.Single.Lesion** |  |  |  | 0.299 |
| Yes | 7 (15.6%) | 24 (23.1%) | 31 (20.8%) |  |
| No | 38 (84.4%) | 80 (76.9%) | 118 (79.2%) |  |
| **Incidental.Multiple.Lesions** |  |  |  | 0.135 |
| Yes | 0 (0.0%) | 5 (4.8%) | 5 (3.4%) |  |
| No | 45 (100.0%) | 99 (95.2%) | 144 (96.6%) |  |
| **Incidental.Other** |  |  |  | 0.210 |
| Yes | 4 (8.9%) | 4 (3.8%) | 8 (5.4%) |  |
| No | 41 (91.1%) | 100 (96.2%) | 141 (94.6%) |  |
| **Incidental.Isolated.Developmental.Venous.Anomaly** |  |  |  | 0.382 |
| Yes | 2 (4.4%) | 2 (1.9%) | 4 (2.7%) |  |
| No | 43 (95.6%) | 102 (98.1%) | 145 (97.3%) |  |
| **Incidental.Infratentorial.Old.Stroke** |  |  |  | 0.100 |
| Yes | 0 (0.0%) | 6 (5.8%) | 6 (4.0%) |  |
| No | 45 (100.0%) | 98 (94.2%) | 143 (96.0%) |  |
| **Incidental.Infratentorial.Encephalomalacia** |  |  |  | 0.509 |
| Yes | 0 (0.0%) | 1 (1.0%) | 1 (0.7%) |  |
| No | 45 (100.0%) | 103 (99.0%) | 148 (99.3%) |  |
| **Incidental.Non.Arachnoid.Cyst** |  |  |  | 0.347 |
| Yes | 1 (2.2%) | 6 (5.8%) | 7 (4.7%) |  |
| No | 44 (97.8%) | 98 (94.2%) | 142 (95.3%) |  |
| **Incidental.Enlarged.Or.Asymmetric.Ventricles** |  |  |  | 0.182 |
| Yes | 0 (0.0%) | 4 (3.8%) | 4 (2.7%) |  |
| No | 45 (100.0%) | 100 (96.2%) | 145 (97.3%) |  |
| **Incidental.Cavum.Septum.Or.Vergae** |  |  |  | 0.349 |
| Yes | 0 (0.0%) | 2 (1.9%) | 2 (1.3%) |  |
| No | 45 (100.0%) | 102 (98.1%) | 147 (98.7%) |  |
| **Incidental.Dilated.Perivascular.Spaces** |  |  |  | 0.250 |
| Yes | 0 (0.0%) | 3 (2.9%) | 3 (2.0%) |  |
| No | 45 (100.0%) | 101 (97.1%) | 146 (98.0%) |  |

Single lesions of significance in epilepsy were found in 11% of patients, but also in 7% of controls. Neoplasms were observed in 4% of patients and no controls (p = 0.18). The next most common epilepsy-related finding was supratentorial encephalomalacias (3% of patients and 4% of controls) and malformations of cortical development (3% of patients and no controls). White matter hyperintensities of unknown significance in epilepsy were observed in 49% of patients, compared to 24% of controls (p = 0.08). Similarly, single lesions of unknown significance were found in 49% of epilepsy patients and 33% of controls (p = 0.08), while 14% of patients and 2% of controls had multiple unknown lesions (p = 0.03). Asymmetry of the amygdala and hippocampus was identified in 4% and 2% of patients (respectively) and in 0% of controls (p = 0.13, p = 0.35). Single lesions not thought to be related to epilepsy (i.e., incidental) were observed in 23% of patients and 16% of controls (p = 0.3), and 5% of patients (no controls) had multiple incidental lesions (p = 0.13). Six percent of patients (no controls) had an infratentorial old stroke, and 4% (no controls) had enlarged or asymmetric ventricles (p = 0.18).

### Quantitative structural MRI

After correction for multiple comparisons, there were no significant differences in subcortical volumes, regional cortical thickness, or cerebellar volumes between patients and controls (Supplementary Table 2). Given that we expected morphometric alterations in the early stages of epilepsy to be subtle, we also explored differences without FDR correction, as the direction and pattern of findings provide information about overall trends that may become more pronounced with disease progression. Results indicated no differences in subcortical or cerebellar volume but decreased cortical thickness predominantly in bilateral frontal regions. In the left hemisphere, this included precentral (t=2.429, p=0.016), rostral middle frontal (t=2.117, p=0.036), and superior frontal (t=2.00, p=0.047) cortex. In the right hemisphere, caudal middle frontal (t=2.287, p=0.024), paracentral (t=2.015, p=0.046), pars opercularis (t=2.045, p=0.043), rostral middle frontal (t=2.207, p=0.029), and superior frontal (t=2.541, p=0.012) cortical regions were thinner in patients. Figure 3A shows the Cohen’s *d* effect sizes of regional volume and cortical thickness differences and illustrates reduced cortical thickness predominantly in hemispheric frontal lobe regions in patients compared to controls.

**Figure 3.**
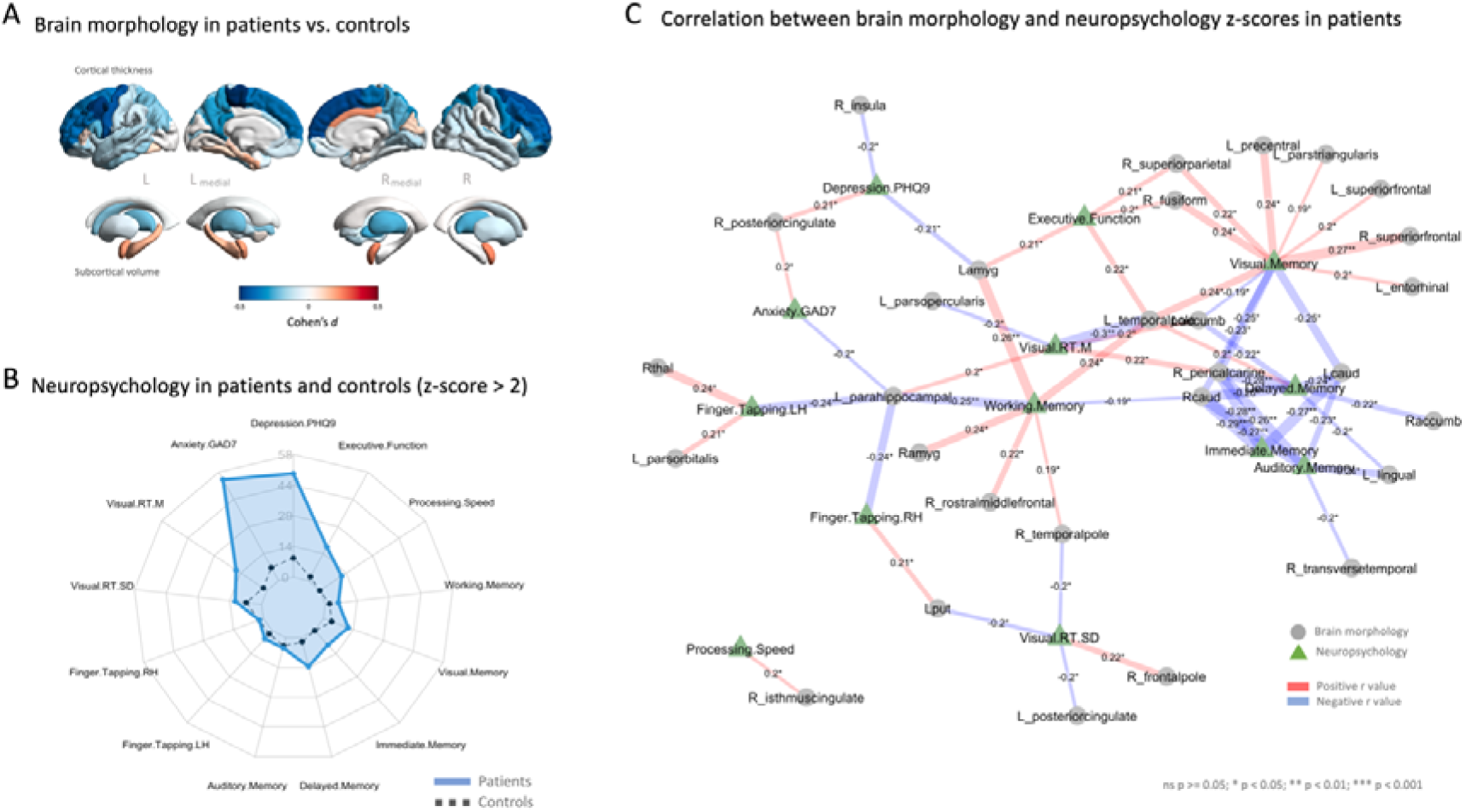
Quantitative MRI and neuropsychological differences between patients and controls. A. MRI brain morphometrics. Differences displayed as Cohen’s *d* in regional cortical thickness (top) and subcortical volumes (bottom) in patients with NDE compared to healthy controls. Blue indicates larger values in controls, red indicates larger values in patients. B. Neuropsychology radar plot indicating the percentage of patients with NDE with z-scores greater than 2 SD for each neuropsychological feature (blue) compared to healthy controls (black). C. Significant correlations (p < 0.05 uncorrected) between neuropsychological and mood measure z-scores (green triangles) and brain morphometric z-scores (grey circles) in patients. Edge colour shows negative (blue) and positive (red) correlations, while edge size indicates the strength of the relationship. Pearson correlation coefficient and associated significance level (*) are shown at the centre of each connection.

### Neuropsychological findings

Table 3 presents the neuropsychological findings in our cohort of patients and controls. Patients had significantly lower scores on tests of executive function, processing speed, visual memory, delayed memory, and immediate memory compared to controls. Patients also had significantly higher scores on measures of depression and anxiety. The discrepancy between patients and controls on tests of depression and anxiety exceeded that of executive function, processing speed and memory. This difference was reflected in the proportion of patients calculated to exceed two SD on individual tests, as illustrated in Figure 3B. 49% and 54% of patients had scores on tests of depression and anxiety greater than two SD compared to 9% and 7% in controls, respectively. This contrasted to 18%, 12%, 12%, 14%, and 9% of patients and 2%, 0%, 4%, 2%, and 0% of controls in tasks of executive function, processing speed, visual memory, delayed memory, and immediate memory, respectively. The relationship between domains of neuropsychological performance found to be impaired in patients and morphometric MRI data are presented in Figure 3C. The direction of the association (red = positive correlation, blue = negative correlation) and strength of association (size of connections, level of significance) are indicated for all significantly related variables.

**Table 3.**
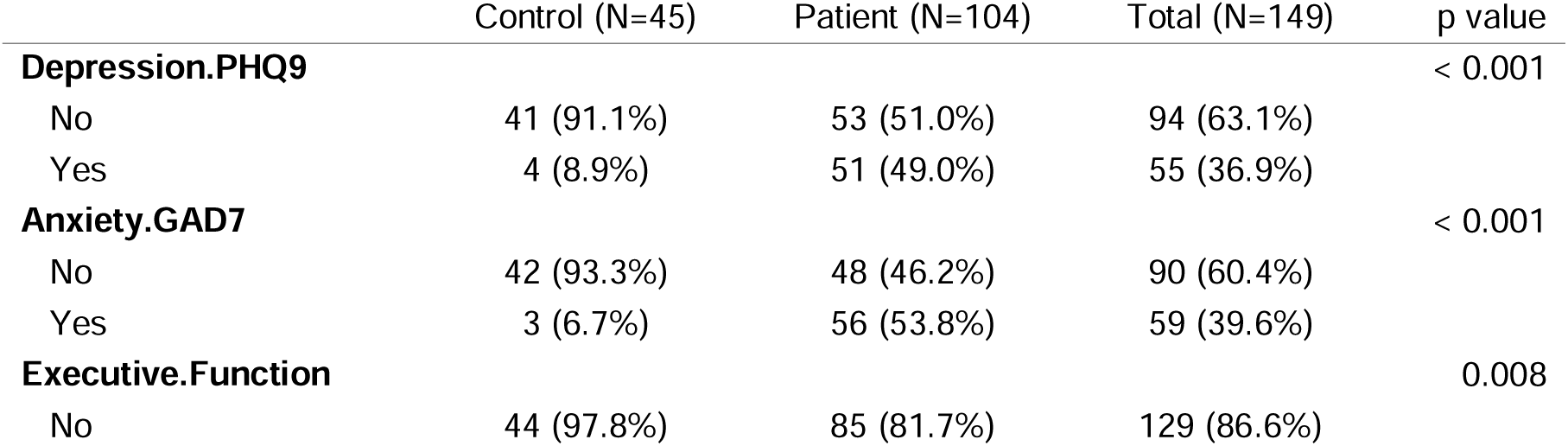
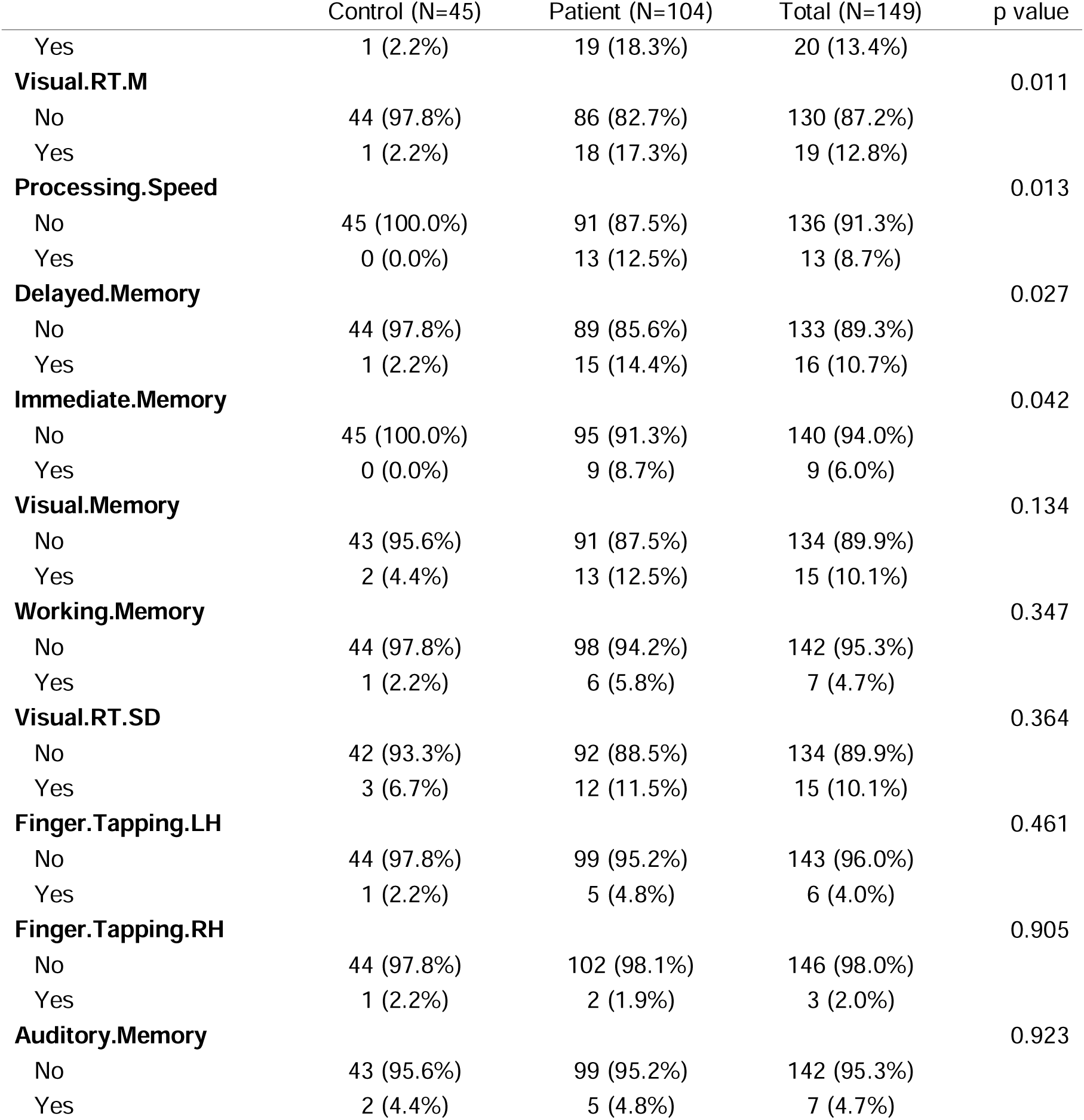
Neuropsychological characteristics of patients with NDE and controls: z-score > 2.

## Discussion

We report baseline assessment data in an adult cohort of patients with focal NDE and matched healthy controls who received standardised neuroimaging, EEG, and neuropsychological evaluation. The novelty in our study includes prospective multi-modal research grade data acquisition and the inclusion of healthy controls enabling us to directly compare neuroradiological and neuropsychological findings using a systematic approach. Our neuroradiological findings indicate a unexpectantly high number of positive MRI findings in patients and controls, and a greater proportion of lesions with unknown relevance to epilepsy in patients, particularly focal white matter hyperintensities. Quantitative neuroimaging data suggested moderately reduced cortical thickness in the frontal lobe and thalamus in patients compared to controls. Neuropsychological data indicated significantly greater depression and anxiety scores in patients compared to controls, and moderate impairments in executive function, processing speed and delayed and immediate memory, impairments that were related to cortical and subcortical morphometric data. Finally, we observed striking changes in seizure outcome status at six-month intervals in the patients who had multiple follow up seizure data.

### Neuroimaging findings

A recent study reported that 59.3% of adolescents and adults with focal NDE were MRI-negative, 18.7% had lesions that were considered epilepsy-related, 17% had abnormalities with unknown relationship to epilepsy, and 5% had incidental findings.^19^ Our findings were 38%, 12%, 49% and 23%, respectively. The largest difference was a higher proportion of patients with abnormalities with unknown relationship to epilepsy, which was driven by a high number of patients with white matter alterations particularly conspicuous on T2-FLAIR images. One significant strength of our study was that we also collected the same imaging data for neurological healthy controls to which our patients could be compared. The number of patients with white matter abnormalities was proportionally greater than controls, which suggests that these non-specific findings could have relevance for adult-onset epilepsy. White matter hyperintensities represent macrostructural brain damage associated with multiple underlying causes and implications,^34, 35^ and should be considered potentially clinically significant. A leading cause of focal white matter lesions is small vessel ischemia, which is has been associated with focal onset seizures in some studies.^36, 37^ The higher incidence of white matter changes in patients could not be attributed to an aging cohort given that our controls were slightly older than our patients. We are unsure as to the reason for the discrepancy in the reported proportion of patients with white matter abnormalities between our study and the aforementioned previous study,^19^ but this could be due to methodological factors such as our use of identical MRI sequences, a single high-end 3 T scanner and a single epilepsy-experienced neuroradiologist for evaluation all patients and controls. The Siemens Magnetom Prisma 3 T MRI system used in our study offers excellent diagnostic sensitivity of white matter hyperintensities on 3D T2-FLAIR images.

Our results are consistent with previous reports of a low proportion of adults with NDE presenting with known epileptogenic lesions, including hippocampal sclerosis and malformations of cortical development.^19, 38^ We did not observe a single case with clear hippocampal sclerosis (although hippocampal asymmetry was reported in one patient). Unlike previously,^19^ we classified supratentorial arachnoid cysts as related to epilepsy (unless there was clear evidence to suggest an alternative cause) as these lesions have been associated with epilepsy in previous studies.^21–23^ However, the inclusion of these lesions as potentially epileptogenic did not increase the incidence of epilepsy-related lesions in our cohort compared to the previous study.^19^

After stringent correction for multiple comparisons, we report no statistically significant changes in regional compartmental brain volume in patients with NDE compared to controls, although moderate effect sizes were observed for bilateral thalamic volume reduction and especially frontal lobe cortical thinning. There are few studies employing quantitative MRI approaches in NDE. We have previously reported thalamic volume reduction in an independent heterogenous group of patients with non-lesional focal NDE using manual MRI measurements,^39^ and thalamic atrophy is a known pathoanatomical feature of longstanding focal and generalised epilepsy.^40^ Similar to our work, other studies have reported no significant changes in hippocampal volume in focal NDE.^41, 42^ Some^42^ but not others^43^ have reported cerebellar volume changes at the time of diagnosis. There are fewer studies examining cortical alterations in NDE, although one longitudinal study in patients with established focal epilepsy reported that the most pronounced ‘progressive’ cortical degeneration was observed in the first five years after the onset of seizures.^44^ Despite that our cohort may be clinically heterogeneous, there is a clear pattern of grey matter changes in the frontal lobe that may be considered as subtle atrophy in the early stages of focal epilepsy. Gross brain alterations are more subtle in focal NDE, which is likely due to a combination of heterogenous groups of patients (compared to, for example, refractory temporal lobe epilepsy in whom clear patterns of gross brain alterations are observed) and limited insights into architectural and network abnormalities that are not amenable to investigation using MRI data acquired in context of routine clinical care. However, with respect to heterogeneous groups of patients, imaging markers of treatment outcome could be common across the different foci in NDE given that; (i) widespread alterations in network structure can give rise to a clearly localised focal onset in one brain region;^45^ (ii) particular anatomical circuits act as critical modulators of seizure generation and propagation, and seizure activity does not spread diffusely throughout the brain but propagates along specific anatomical pathways, regardless of the localisation of the brain insult;^46, 47^ (iii) pathological structural connectivity causes disturbances to common large scale functional brain networks regardless of the localisation of the epileptogenic zone in patients with refractory focal epilepsy;^48^ (iv) particular deep brain regions - such as the thalamus - that play a crucial role in the clinical manifestation of seizures in the epilepsies,^49^ and anatomically support widespread distributed cortico-subcortical networks,^50^ are structurally and physiologically abnormal in both hemispheres in patients with longstanding focal and generalised epilepsy disorders.^40^ We propose that some brain alterations in epilepsy are established by the time of first seizure and may result from genetic factors, developmental processes, occult insult, or other mechanisms of epileptogenesis.

### Neuropsychological findings

It is well established that chronic epilepsy is associated with cognitive and psychiatric deficits that are dependent on the semiology, aetiology, and phenotype of epilepsy disorder,^51, 52^ and that deficits may progressively deteriorate as uncontrolled seizures accumulate.^13, 52, 53^ It is equally well established that cognitive and affective deficits are present in drug naïve patients with NDE, particularly in the domains of memory, sustained attention, executive functioning, mental flexibility and psychomotor speed.^5–10^ Our work supports these findings. A previous study reported that impairments in attention and executive functions were seen in 49.4% and memory deficits in 47.8% of patients with drug naïve patients with NDE, and recommended cognitive screening in new-onset epilepsies,^54^ which is not currently embedded in clinical evaluation pathways. Most patients underreport cognitive problems in the early stages of epilepsy as the new onset of seizures cause the greatest concern, but early neuropsychological monitoring could improve individual medical care throughout a lifetime lived with epilepsy.^54, 55^

In children with new onset epilepsy, mild diffuse cognitive impairment was not related to MRI morphometric total cerebral or lobar tissue volumes.^56^ This may suggest that the neurobiological substrate of cognitive impairment in NDE exists beyond what can be inferred from examining gross brain morphology. With respect to neuroradiological findings, it has been previously reported that lesional NDE is associated with more deficits in attention and executive functions compared to non-lesional NDE and worse memory performance was related to generalised tonic-clonic seizures.^54^ We report significant correlations between various neuropsychological and mood measures and brain morphometric variables (Figure 3C), which require further investigation. To our knowledge there are no other studies examining the relationship between neuropsychological performance and quantitative morphometrics in NDE and therefore future work is required to examine our preliminary findings. Longitudinal imaging and neuropsychological studies will be important in determining whether progressive deterioration in cognition^11^ are related to pathological progressive brain changes through the early stages of epilepsy. Moreover, given that psychiatric problems including depression may antedate the diagnosis of epilepsy in adults,^57, 58^ it will be important to monitor brain-psychiatric coupling throughout early epilepsy.

### Future work

Given that neuroradiological and volumetric MRI data shed limited insights into seizure and cognitive status in NDE, it is important to phenotype and stratify patients using imaging methods sensitive to the microstructural and network architecture of the brain. It is likely that imaging markers of pharmacoresistance will be microstructural, functional, or metabolic.^39^ Prognostication of patients with refractory epilepsy has been reasonably successful using brain connectivity and network approaches,^14–16^ and therefore the utility of similar approaches to predict pharmacoresistance and shed light on the mechanistic underpinnings of cognitive dysfunction need to be explored. We are in the novel position of acquiring imaging data amenable to investigation of microstructural brain architecture and structural and functional brain networks in patients with NDE and will seek to determine whether these facets of brain organisation have mechanistic and clinical utility in the early stages of epilepsy. Furthermore, given the large proportion of patients with white matter hyperintense lesions, future work should quantitatively examine the relationship between these lesions and neuropsychological deficits in our sample, particularly given the previously reported association between the two in patients with different neurological disorders.^35^

## Conclusion

The prevalence of MRI negative cases in patients with focal NDE is lower than what previously thought. A significant contributor to this is the prevalence of white matter hyperintensities and the relationship between these and onset of focal epilepsy requires further investigation. Heterogeneous cohorts of patients with focal NDE do not present with strong group-wise changes in brain morphometry, but moderate size alterations are present in the frontal lobe and thalamus. Neuropsychological and mood impairments are present at diagnosis, some of which are related to gross morphometric brain alterations, which suggests that cognitive and psychiatric assessment should be considered in the early stages of epilepsy, which could support cognitive rehabilitation strategies.

## Data Availability

All data produced in the present study are available upon reasonable request to the authors

## Acknowledgements

This work was funded by a UK Medical Research Council (MRC) research grant (MR/S00355X/1). We thank all health care professionals at the Walton Centre and Salford Royal NHS Foundation Trusts for helping to recruit patients into this study.

